# Impact of Calcium Cracks after Balloon Angioplasty in Patients with Complex Calcified Coronary Plaque ∼The Results of the OCT-CALC Registry∼

**DOI:** 10.1101/2024.02.17.24302734

**Authors:** Nobuhiko Maejima, Tsutomu Endo, Takashi Ashikaga, Taishi Yonetsu, Kazuhiro Ashida, Hiroshi Ohira, Takahiko Kiyooka, Tomohiko Shigemasa, Gaku Nakazawa, Yuji Ikari, Akihiro Hata, Tetsuya Tobaru, Itaru Takamizawa, Teruyasu Sugano, Ichiro Michishita, Kouji Yamamoto, Toshiro Shinke, Ken Kozuma, Yasuhiro Honda, Kiyoshi Hibi

**Author notes:** Correspondence to: Kiyoshi Hibi, M.D. Department of Cardiology, Yokohama City University Graduate School of Medicine 3-9 Fukuura, Kanazawa-ku, Yokohama 236-0004, Japan.

## Abstract

**Background:** Target lesion calcification is known to influence the percutaneous coronary intervention (PCI) outcomes. Calcium cracks as assessed by optical coherence tomography (OCT) after balloon angioplasty were associated with a larger stent area and a larger lumen gain after PCI for the lesions with moderate to severe calcification, although clinical outcomes in those patients remain unclear. This study aimed to assess the impact of calcium cracks after balloon angioplasty on the PCI results as well as the long-term clinical outcomes by multicenter OCT-guided PCI registry.

**Methods:** We formed a prospective, multicenter registry that include 22 sites from Japan and Korea that enrolled 268 patients who underwent PCI to the lesion with moderate to severe calcification on angiogram. Balloon dilatation and subsequent drug elution stent (DES) implantation were performed with OCT guidance in every case. Lesion modification with rotational atherectomy was performed before balloon dilatation if needed. Serial OCT images just before and after balloon angioplasty, and after stent implantation were analyzed at 1-mm intervals by an independent core laboratory. The incidence of calcium cracks after balloon angioplasty was assessed at the minimal lumen area (MLA) site by OCT. By protocol, follow-up angiography was performed 10 months after PCI (in 85.5% of patients), and both baseline and follow-up angiograms were analyzed by an angiographic core laboratory. The primary endpoint was the relationship between calcium crack after balloon angioplasty and stent expansion. The secondary endpoint was target vessel failure (TVF) at 1 year, defined as a composite of cardiac death, target vessel-related myocardial infarction, and target vessel revascularization.

**Results:** A total of 242 patients were analyzed. Of these, OCT analysis was performed in 147 patients with a complete OCT data set. Calcium cracks were observed in 28 patients (19%) at the MLA site. The percent stent expansion was greater in lesions with calcium crack than those without (99±26 % vs. 91±18 %, p=0.039). In 229 patients who underwent clinical follow-up at 1 year, TVF occurred in 23 patients (10.0%). In 139 patients in whom both OCT analysis and 1-year clinical follow-up was performed, the incidence of TVF was similar between patients with and without crack formation (11 % vs. 13 %, p=1.00). Angiographic sub-analysis with both baseline and 10-months follow-up was performed in 124 patients. Acute lumen gain, as well as late lumens loss, were greater in patients with calcium crack than those without (1.39±0.55 mm vs. 1.15±0.48 mm, p=0.037; 0.51±0.67 mm vs. 0.12±0.51 mm, p=0.0095, respectively), resulting in similar net lumen gains between the 2 groups.

**Conclusion:** The OCT-guided PCI strategy demonstrated acceptable acute and 1-year clinical outcomes. The presence of calcium cracks after balloon angioplasty may have a potential impact on acute results after DES implantation in calcified lesions. However, its impact may be attenuated by late lumen loss at 10-months follow-up.

## Introduction

Calcified lesions are the most challenging subset of lesions during percutaneous coronary intervention (PCI). They adversely interfered with the ability to cross or dilate a coronary stenosis with PCI devices such as balloons or stents, leading to poor acute outcomes including stent thrombosis (1, 2). Despite recent advancements in drug eluting stents (DES), target lesion failure after PCI of calcified lesions remains high, ranging from 10.7 to 20.9% (3–5). Recently, several studies have demonstrated that intravascular ultrasound (IVUS) improved outcomes after PCI (6–8), especially in complex lesions (9, 10). However, the IVUS-guided strategy failed to show a beneficial effect on cardiovascular events in patients with calcified lesions (11). Optical coherence tomography (OCT) is an optical analogue of IVUS that can visualize calcium components with less signal attenuation (12), enabling full delineation of superficial calcium components in the arterial wall (13). A potential application of OCT is to provide detailed information about the structural change of plaque components during a PCI procedure. We and other researchers have previously shown that calcium cracks as assessed by OCT after balloon angioplasty were associated with a larger stent area and a larger lumen gain after PCI in patients with calcified lesions (14–16). However, most studies are single-center observational studies with a small number of patients, and the impact of calcium cracks on clinical outcomes in lesions with moderate to severe calcification remains unclear. Therefore, we conducted a prospective multicenter registry to explore the factors associated with calcium crack after balloon angioplasty in patients with moderate to severe lesion calcifications and to assess the impact of calcium cracks on the PCI results and long-term clinical outcomes.

## Methods

### Study design and patients

The OCT-CALC (Optical Coherence Tomography assessment for Coronary Artery Lesions with Calcification) registry was a prospective, multicenter registry including 22 sites from Japan and Korea that include both sites capable and not capable of performing rotational atherectomy (RA). The study enrolled patients with de novo calcified lesions in a native coronary artery who underwent OCT-guided PCI. Patients were eligible for enrollment if: 1) they were indicated for PCI using DES with OCT guidance; 2) the lesion calcification was moderate to severe as assessed by angiogram (17); and 3) they provided written informed consent. Exclusion criteria were as follows: 1) a culprit lesion for ST segment elevation myocardial infarction; 2) cardiogenic shock; 3) heart failure; 4) a restenotic lesion or graft lesion; 5) a lesion not suitable for stent implantation; 6) a serum creatinine level >2.0 mg/dl without undergoing hemodialysis; or 7) an ostial lesion. Patients in whom adequate OCT images could not be obtained were also excluded from the analysis.

The protocol was approved by the institutional review board at the Yokohama City University Medical Center and each participating center, and the study was performed in accordance with the principles of the Declaration of Helsinki and the Ethical Guidelines for Medical and Health Research Involving Human Subjects. This trial was registered in the University hospital Medical Information Network (UMIN) Clinical Trial Registry (UMIN 000018636).

### Procedures

Balloon dilatation and subsequent DES implantation were performed with OCT guidance in every case. Lesion modification with RA was allowed before balloon dilatation if needed. Orbital atherectomy or intravascular lithotripsy were not available in Japan during the study period. OCT imaging was performed with either frequency-domain, ILUMIEN OCT imaging system; (Abott Vascular, Westford, MA, USA), or LUNAWAVE; (Terumo Corp., Tokyo, Japan). OCT evaluation was attempted just after successful guidewire crossing and after RA if possible, and OCT pullback after ballooning and after final stent implantation were mandatory. Additional preparation or stent optimization was performed based on the OCT image; however, OCT evaluation after additional procedures was also needed. Optimal medical therapy, including dual antiplatelet therapy, was based on the standard of care. The medication regimen and PCI strategy including DES selection and stent sizing were at the discretion of the attending physician or the PCI operator.

### Quantitative coronary angiographic analysis

Coronary angiography was performed with a frame rate of 30 frames/s. Baseline angiography was performed before PCI and follow up angiography was performed 10 months after PCI. Both angiograms were analyzed by an independent core laboratory (Cardiocore, Tokyo, Japan). Quantitative coronary angiography (QCA) was performed using an automated edge-detection system (QAngio XA, Medis, Leiden, the Netherlands). Binary restenosis was defined as a diameter stenosis >50 % at the follow-up angiogram and was determined in-stent and in-segment (including the stent and segments 5 mm proximal and distal to the stent edge). Acute lumen gain and late lumen loss were calculated as (the post-procedural minimal lumen diameter) – (the baseline minimal lumen diameter), and the (the post-procedural minimal lumen diameter) – (the follow-up minimal lumen diameter), respectively.

### OCT image analysis

Off-line OCT analysis was performed using a validated intravascular image analysis system (echoPlaque 4, Indec Systems, Inc., Santa Clara, CA, USA) at an independent core laboratory (Cardiovascular Core Analysis Laboratory, Stanford University, Stanford, CA, USA) blinded to clinical and angiographic information. Serial OCT images were co-registered using fiducial points as landmarks (e.g., side branches or calcium itself), and qualitative/quantitative analyses were performed in a standard manner (18). In cross-sectional analysis, acute lumen gain was calculated as an increase in minimal lumen area (MLA) from pre-dilatation to post-stenting; percent stent expansion was defined as the ratio of the minimal stent area and the average reference lumen area (%) (19). In volumetric analysis, vessel and stent boundaries were manually traced at 1-mm intervals throughout the target segment with automated interpolated measurements of the remaining frames. Each volume calculated with Simpson’s method was standardized by analyzed length (mm^3^/mm).

Detailed calcium analysis by OCT has been published previously (14). In brief, target lesion calcium deposit was evaluated at pre-dilatation, including calcium arc and minimal calcium thickness (measured at the thinnest part, excluding the outer 30° segment at both edges of the calcium arc). Calcium crack formation was defined as discontinuity of the luminal surface in the calcium component acquired at post-dilatation (Figure 1).

**Figure 1.**
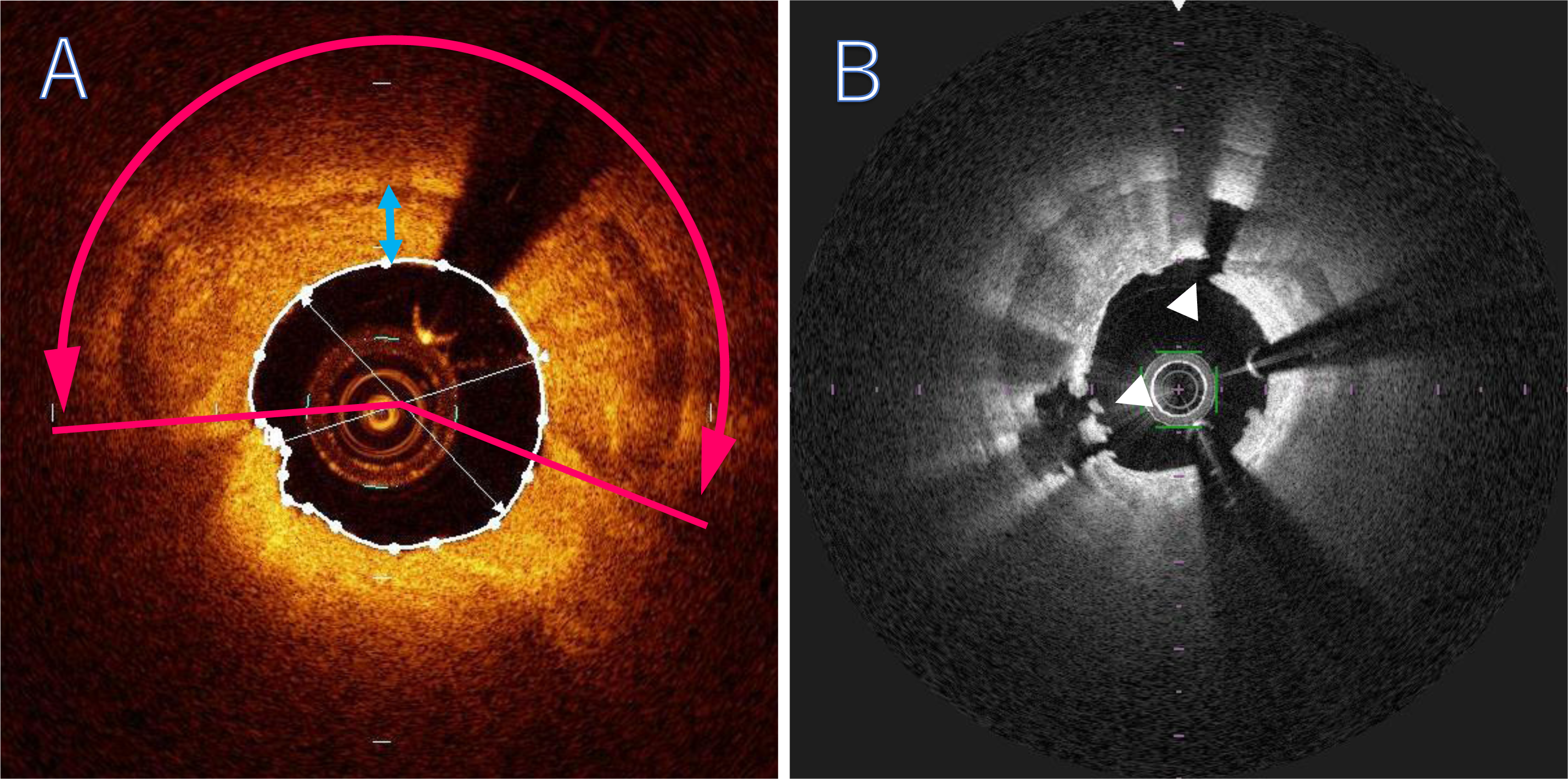
OCT analysis. Calcium arc (double-headed pink arrow), and minimal calcium thickness (double-headed blue arrow) were measured by OCT at pre-dilatation (A). The presence of cracks (arrowheads) was also assessed at post-dilatation (B).

### Study endpoints

The primary endpoint was the relationship between calcium cracks after balloon angioplasty and stent expansion, as assessed by lesion-based analysis. The secondary endpoint was target vessel failure (TVF) at 1 year, defined as a composite of cardiac death, target vessel-related myocardial infarction, and target vessel revascularization. Clinical events were adjudicated by an independent clinical event committee.

### Statistical Analysis

Statistical analysis was performed using JMP 15 software (SAS Institute, Cary, North Carolina). Qualitative data are presented as numbers (%). Normally distributed, continuous variables are expressed as means ± SD and were compared using Student’s t-test. Continuous variables with skewed distributions are expressed as median values (interquartile range) and were compared with the Wilcoxon rank-sum test. To determine the optimal threshold values of the calcium arc and calcium thickness for the prediction of calcium cracks after balloon dilatation, receiver-operating characteristics (ROC) curve analyses were performed. The cutoff point was defined as the greatest sum of the sensitivity and specificity estimates. For all analyses, p values of <0.05 were considered to indicate statistical significance.

## Results

### Study population

A total of 268 patients were enrolled in the OCT-CALC registry. Of these, 26 patients were ineligible for baseline analysis, and thus 242 patients received treatment and comprised the study population (Figure 2). Baseline patient characteristics are summarized in Table 1. One hundred thirteen patients (47%) had diabetes, and 39 patients (16%) received hemodialysis. Procedural characteristics are shown in Table 2. The severity of calcification in the target lesion was moderate in 80 patients (33%) and severe in 162 patients (67%). RA was performed in 83 patients (34%). RA was available in 13 of the 22 study participant centers, and 171 patients (71%) were enrolled from RA-capable centers.

**Figure 2.**
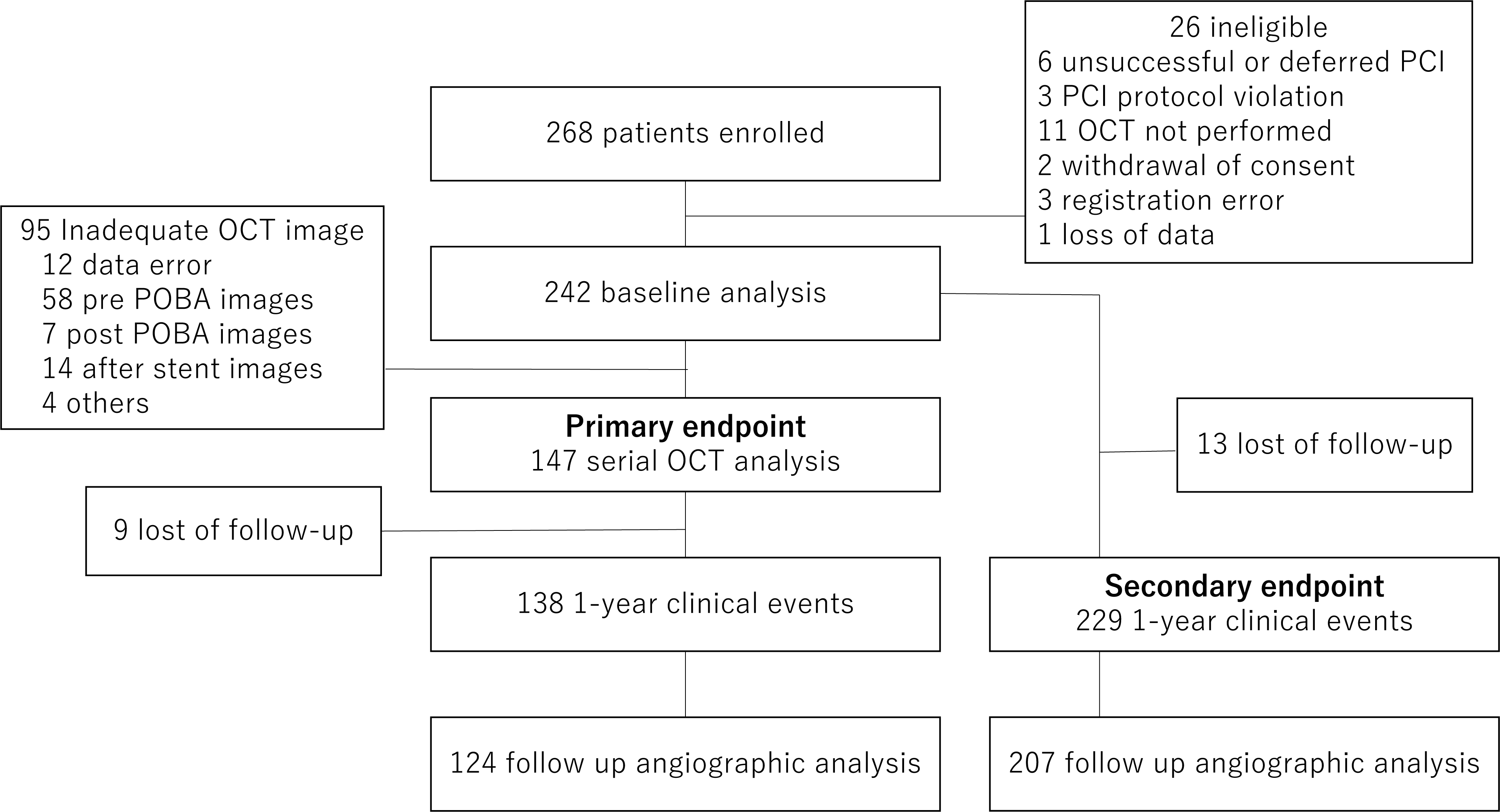
Study flowchart for the Optical Coherence Tomography assessment for Coronary Artery Lesions with Calcification (OCT-CALC) registry.

**Table 1.** Baseline characteristics.

|  | n=242 |
| --- | --- |
| Age, yrs | 72.4±8.8 |
| Male gender (%) | 173 (71%) |
| Coronary risk factors |  |
| Diabetes (%) | 113 (47%) |
| Hypertension (%) | 189 (78%) |
| Dyslipidemia (%) | 153 (63%) |
| Smoking (%) | 102 (42%) |
| Chronic kidney disease (%) | 116 (48%) |
| Hemodialysis (%) | 39 (16%) |
| Clinical presentation |  |
| Acute coronary syndrome | 27 (11%) |
| Stable angina pectoris | 215 (89%) |

Data presented are means ±SD or numbers (%) of patients.

**Table 2.** Procedural characteristics.

|  | n=242 |
| --- | --- |
| Target vessel |  |
| Left anterior descending | 178 (74%) |
| Left circumflex artery | 16 (7%) |
| Right coronary artery | 48 (20%) |
| Lesion calcification |  |
| None/Mild | 0 (0%) |
| Moderate | 80 (33%) |
| Severe | 162 (67%) |
| Procedural characteristics |  |
| Rotablator use | 83 (34%) |
| Number of Rotablator | 1.26±0.44 |
| Final burr size (mm) | 1.61±0.17 |
| Scoring balloon use | 147 (61%) |
| Number of stents | 1.34±0.55 |
| Stent diameter | 3.01±0.43 |
| Total stent length / lesion | 34.2±16.1 |
| Maximal stent pressure (atm) | 13.6±2.9 |
| Post dilation | 172 (71%) |
| Maximal balloon diameter (mm) | 3.24±0.57 |
| Maximal balloon pressure (atm) | 17.4±4.5 |

Data presented are means ± SD or numbers (%) of patients.

A one-year clinical follow-up, and 10-months angiographic follow-up were performed in 229 patients and 207 patients, those were 94.6% and 85.5% of the total population, respectively (Figure 2). One-year clinical events are shown in Table 3. TVF occurred in 23 patients (10.0%). Patients and angiographic lesion characteristics were not associated with TVF (Supplement Table).

**Table 3.** Event at follow-up.

|  |  |
| --- | --- |
| 1-year clinical event (n=229) |  |
| Target vessel failure | 23 (10.0%) |
| All cause death | 12 (5.2%) |
| Cardiac death | 4 (1.7%) |
| Myocardial infarction | 4 (1.7%) |
| Stent Thrombosis | 3 (1.3%) |
| Target lesion revascularization | 11 (4.8%) |
| Target vessel revascularization | 19 (8.3%) |
| 10-months angiographic follow-up (n=207) |  |
| Binary restenosis | 24 (11.6%) |

Data presented are numbers of patients.

The OCT analysis was performed to explore the factors associated with acute PCI results, as well as clinical outcomes. After 95 patients were excluded from OCT analysis, OCT images were analyzed in 147 patients. Of these, 1-year clinical and 10-months angiographic follow-up was performed in 138 patients (93.9%) and 124 patients (84.4%), respectively (Figure 2).

### OCT analysis

The results of the OCT analysis are shown in Table 4. Calcium cracks at the MLA site were observed in 28 patients (19%). When we stratified study participants into 2 groups based on the presence or absence of calcium cracks after balloon dilation at the MLA site, baseline and procedural characteristics were comparable between the 2 groups. The calcium arc was greater and the minimal calcium thickness was thinner in the crack group than in the no-crack group (273, IQR 188-339 degrees, vs. 119, IQR 91-172 degrees, p<0.0001; 0.44±0.25 mm vs. 0.66±0.30 mm, p=0.0008, respectively), while the lumen area before balloon dilatation was similar between the two groups (1.93±0.65 mm^2^ vs. 2.11±1.17 mm^2^, p=0.45). On ROC curve analysis, the optimal cut-off of calcium arc and minimal calcium thickness for the prediction of calcium cracks was >204 degrees (sensitivity, 75%; specificity, 82%; AUC, 0.806; P<0.0001) and <0.53 mm (sensitivity, 71%; specificity, 61%; AUC, 0.700; P=0.0015), respectively (Figure 3). As shown in Table 4, the percent stent expansion was significantly greater in the crack group than in the no-crack group (99±26 % vs 91±18 %, p=0.039), whereas the final stent area was not statistically different between the two groups (6.21±1.90 mm^2^ vs. 5.84±1.90 mm^2^, p=0.36). Lumen gain was also greater in the crack group than in the no-crack group at the MLA site (4.89±1.95 mm^2^ vs. 3.95±1.57 mm^2^, p=0.007). The malapposed area was greater in the crack group than in the no-crack group (0.70±0.53 mm^2^ vs 0.33±0.32 mm^2^, p<0.0001).

**Figure 3.**
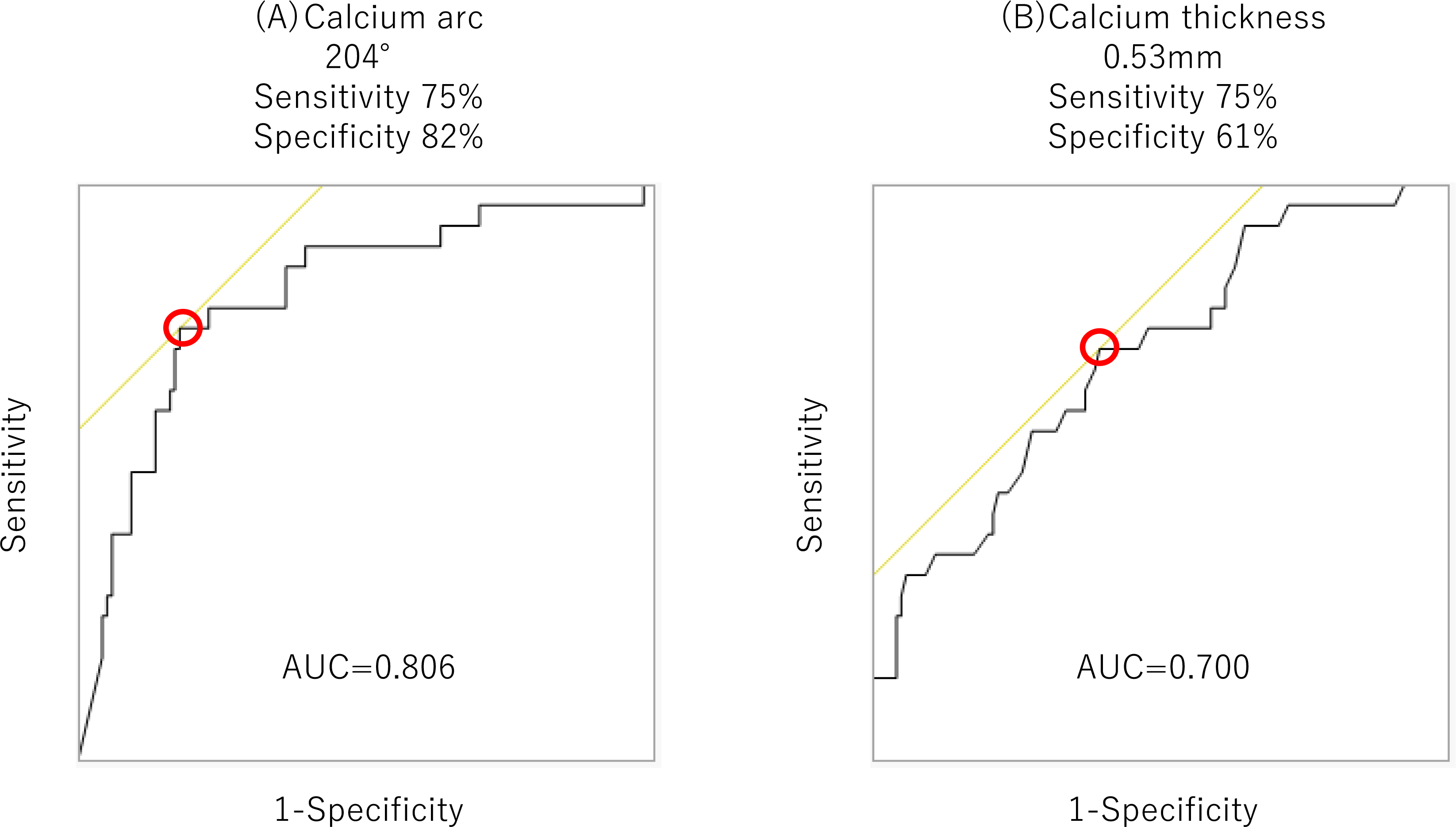
Receiver operating characteristic curve analysis for predicting calcium cracks after balloon angioplasty. The optimal threshold for predicting calcium cracks was >204 degrees (A) and <0.53 mm (B). Red circles indicate optimal threshold. AUC = area under the curve.

**Table 4.** OCT analysis.

|  | Crack<br>n=28 | No-crack<br>n=119 | p Value |
| --- | --- | --- | --- |
| OCT before ballooning |  |  |  |
| Calcium arc (degree) | 273 (188-339) | 119 (91-172) | <0.0001 |
| Minimal calcium thickness (mm) | 0.44±0.25 | 0.66±0.30 | 0.0008 |
| Lumen area (mm<sup>2</sup>) | 1.93±0.65 | 2.11±1.17 | 0.45 |
| OCT after stenting (MLA site analysis) |  |  |  |
| Stent area (mm<sup>2</sup>) | 6.21±1.90 | 5.84±1.90 | 0.36 |
| Percent stent expansion (%) | 99±26 | 91±18 | 0.039 |
| Lumen area (mm<sup>2</sup>) | 6.83±2.10 | 6.06±1.94 | 0.066 |
| Malapposed area (mm<sup>2</sup>) | 0.70±0.53 | 0.33±0.32 | <0.0001 |
| Intimal area (mm<sup>2</sup>) | 0.09±0.12 | 0.12±0.13 | 0.28 |
| Lumen gain (mm<sup>2</sup>) | 4.89±1.95 | 3.95±1.57 | 0.007 |
| OCT after stenting |  |  |  |
| Minimal stent area (mm<sup>2</sup>) | 5.10±1.61 | 5.20±1.75 | 0.79 |
| OCT after stenting (volumetric analysis) |  |  |  |
| Stent volume (mm<sup>3</sup>/mm) | 6.50±0.31 | 6.31±0.17 | 0.63 |
| Lumen volume (mm<sup>3</sup>/mm) | 7.09±0.37 | 6.65±0.184 | 0.28 |
| Malapposed volume (mm<sup>3</sup>/mm) | 0.65±0.05 | 0.43±0.02 | <0.0001 |
| Intimal volume (mm<sup>3</sup>/mm) | 0.06±0.01 | 0.09±0.00 | 0.04 |
| Lumen volume gain (mm<sup>3</sup>/mm) | 2.73±1.60 | 2.70±1.05 | 0.92 |

Data presented are means ± SD or median value (interquartile range).
OCT = optical coherence tomography, MLA = minimal lumen area,

### Angiographic sub-analysis

The quantitative angiographic findings in 147 patients with OCT analysis are listed in Table 5. Lesion length, reference diameter, minimal lumen diameter, and diameter stenosis were comparable between the 2 groups at baseline. In 124 patients who underwent angiographic and clinical follow-up, binary restenosis was observed in 15 patients (12%). Acute lumen gain and late lumen loss were greater in the crack group than in the no-crack group (1.39±0.55 mm vs. 1.15±0.48 mm, p=0.037; 0.51±0.67 mm vs. 0.12±0.51 mm, p=0.0095, respectively), leading to similar net lumen gain (0.91±0.62 mm vs. 0.99±0.51 mm, p=0.57). We also explored the relationship between malapposition and late lumen loss. The malapposed area was positively correlated with late lumen loss (R²=0.078, p=0.0016). The rate of binary restenosis and the incidence of TVF were similar between the two groups, as well as patients and procedural characteristics (Tables 5 and 6).

**Table 5.**
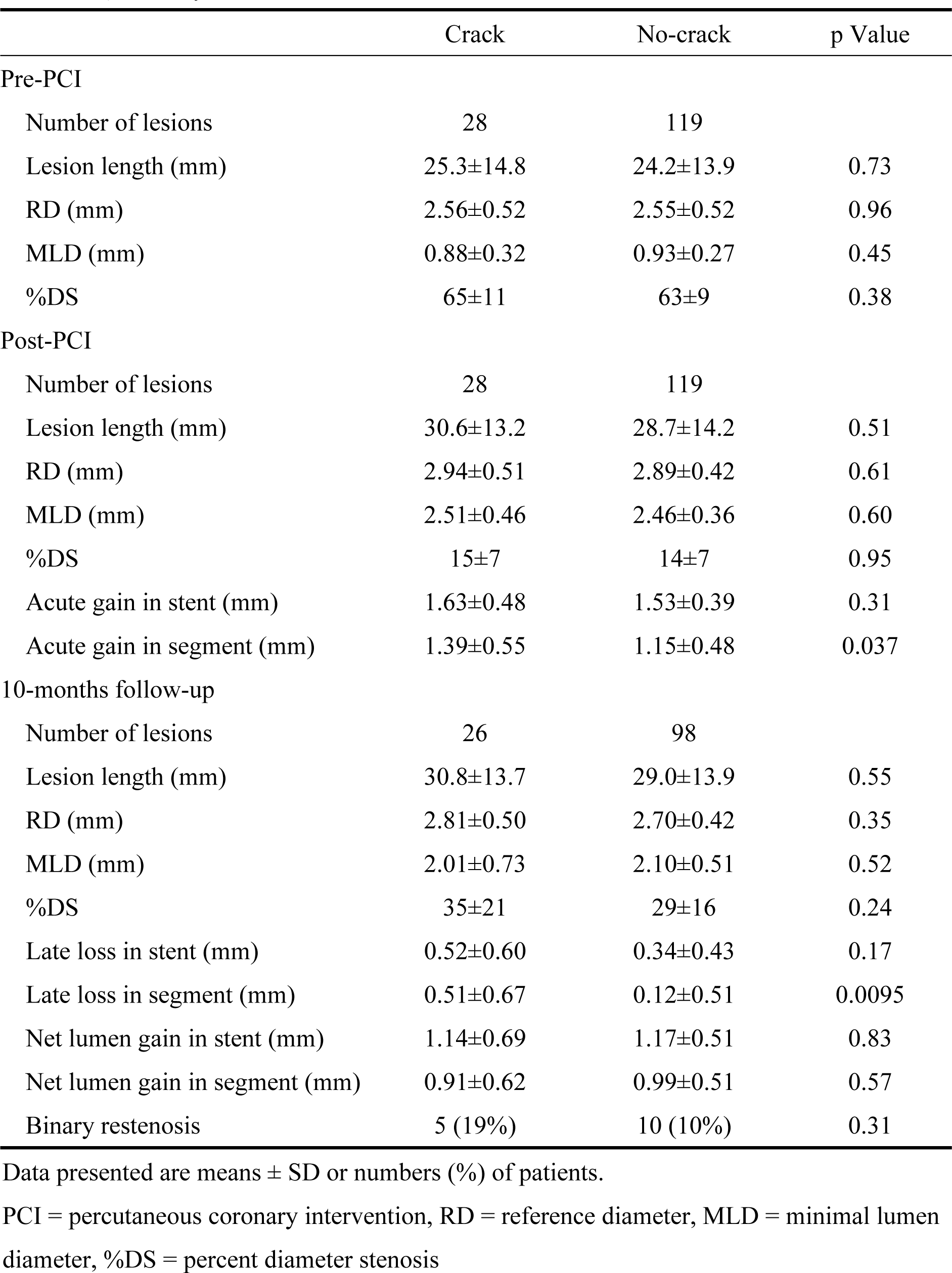
QCA analysis.

**Table 6.** Clinical event.

|  | Crack<br>(n=28) | No-crack<br>(n=110) | p Value |
| --- | --- | --- | --- |
| Patient characteristics |  |  |  |
| Age, yrs | 73±8 | 73±9 | 0.66 |
| Male gender (%) | 20 (71%) | 76 (69%) | 1.00 |
| Diabetes (%) | 13 (46%) | 54 (49%) | 0.84 |
| Hypertension (%) | 23 (82%) | 84 (76%) | 0.62 |
| Dyslipidemia (%) | 18 (64%) | 69 (63%) | 1.00 |
| Smoking (%) | 10 (36%) | 41 (37%) | 1.00 |
| Chronic kidney disease (%) | 16 (57%) | 51 (46%) | 0.40 |
| Hemodialysis (%) | 6 (21%) | 21 (19%) | 0.79 |
| Acute coronary syndrome (%) | 4 (14%) | 7 (6%) | 0.23 |
| Procedural characteristics |  |  |  |
| Severe calcification | 22 (79%) | 74 (67%) | 0.36 |
| Rotablator use | 11 (39%) | 42 (38%) | 1.00 |
| Scoring balloon use | 16 (57%) | 66 (60%) | 0.83 |
| Clinical event |  |  |  |
| Target vessel failure | 2 (8%) | 9 (8%) | 1.00 |
| All cause death | 1 (4%) | 7 (6%) | 1.00 |
| Cardiac death | 0 (0%) | 1 (1%) | 1.00 |
| Myocardial infarction | 0 (0%) | 1 (1%) | 1.00 |
| Stent Thrombosis | 0 (0%) | 0 (0%) | 1.00 |
| Target vessel revascularization | 2 (8%) | 8 (7%) | 1.00 |

Data presented are means ±SD or numbers (%) of patients.

## Discussion

This is a prospective, multicenter registry that enrolled patients who underwent OCT-guided PCI to calcified lesions to evaluate the impact of calcium cracks after balloon dilatation on the acute results of PCI, as well as 1-year clinical outcomes. The primary findings of the present study were that, 1) 1-year clinical events were acceptable in the OCT-guided PCI multicenter registry for lesions with moderate to severe calcification; 2) calcium cracks at the MLA site were associated with greater stent expansion although its impact was diminished by the greater late loss during follow-up; and 3) the impact of calcium cracks were modest with regard to the incidence of TVF at 1 year.

The recent development of DES has substantially ameliorated PCI outcomes and thus, is frequently used in complex lesions including calcified stenosis. However, it is reported that target lesion calcification was associated with stent underexpansion (20), which is a known risk factor for stent restenosis and stent thrombosis (21, 22). Even among the newer generation DES, coronary calcification was associated with unfavorable clinical outcomes after PCI (3–5). In previous reports, the major adverse composite endpoints at 1 year in lesions with moderate to severe calcification was reported to be 10.7—20.9%. Recently, several studies have demonstrated the beneficial effects of IVUS-guided strategies on PCI outcomes especially for complex lesions (6–9). However, its impact on PCI outcomes in calcified lesions remains unclear (11). OCT is a unique imaging modality that can evaluate calcium thickness in vivo. Recently, clinical outcome following OCT-guided PCI has been reported to be equivalent to those after IVUS-guided PCI (23). To the best of our knowledge, this is the first report exploring the impact of OCT-guided PCI on clinical outcomes in patients with moderate to severe lesion calcification. In this study, TVF at 1year was 10.0%, which seemed acceptable compared to other studies. This favorable result may have been facilitated by the study protocol, as OCT imaging before stent implantation was mandatory and additional procedures after OCT imaging were possible. The incidence of crack formation in the calcium might have been high despite the severity of lesion calcification. Indeed, the minimum stent area (MSA) was 5.18±1.72 mm^2^, and 99 lesions (67%) achieved an MSA greater than 4.5 mm^2^ after PCI, which was the recommended goal as an optimal endpoint (24).

We previously reported that calcium cracks were associated with a larger final stent area in 625 segments from 37 lesions that were treated with rotational atherectomy (14). This single-center study had several limitations including 1) a small number of patients, 2) all patients being treated with rotational atherectomy, 3) no patients received dilatation with a scoring balloon, and 4) the threshold to predict calcium crack was based on segment-based analysis. Segment-based analysis may have a potential risk for bias, as a single longitudinally distributed crack could be analyzed in several consecutive segments. The present study was a prospectively designed multicenter study with various strategies. We performed lesion-based analysis in the present study to minimize the effect of interactions between calcium fragility and longitudinal distributions of calcified plaque. In this study, the percent stent expansion was greater in the crack group than in the no-crack group, which confirms the importance of crack formation at the MLA site in lesions with moderate to severe calcification. We also evaluated the impact of calcium severity on the threshold for calcium cracks after balloon dilation. From the present study, the optimal cutoff value to predict calcium cracks was 204 degrees for the calcium arc and 0.53 mm for the minimal calcium thickness. Several studies have demonstrated optimal values to predict calcium crack (14, 15, 25). Kubo et al. reported the median calcium fracture thickness to be 0.45mm from a single-center retrospective registry (15). They assessed the median calcium thickness of the fracture site after stent implantation and did not provide a threshold to predict calcium crack. Our previous single-center observational study showed that optimal cut-off to predict calcium cracks was 227 degrees for the calcium arc and 0.67 mm for the minimal calcium thickness in lesions requiring rotational atherectomy (14), indicating that this population may have had more complex lesion characteristics than that of the Kubo et al. study. Additionally, Fujino et al. reported that the optimal cut-off to predict calcium cracks was 0.24 mm in patients who had not undergone an atherectomy procedure (25). The difference between our study and that of Fujino et al. may be attributed to heterogeneity in the patient population and plaque modification strategies. Fujino et al. retrospectively studied patients treated with balloon angioplasty only before stent implantation, regardless of the degree of calcification. In fact, calcium fracture was observed in only 2% of lesions with a maximum calcium angle <180 degrees, and 76% of the total population had a calcium angle <180 degrees.

Clinical outcomes were similar between lesions with and without cracks. We excluded patients in whom the OCT catheter could not cross the lesion before balloon dilation because OCT images before ballooning were necessary to detect the MLA site for the lesion-based analysis. As result, the impact of calcium cracks on the primary outcome was evaluated with only 147 patients, which may be underpowered to prove the beneficial effect of calcium crack.

Although the final stent expansion was greater in the crack group, the beneficial effect of crack formation on acute results may be attenuated by the healing response after procedural injury and thereby influence the 1-year clinical outcomes. From QCA analyses, greater acute lumen gain was compensated by greater late loss in the crack group, resulting in similar angiographic results at 10 months. Although calcium crack may confer beneficial effects regarding stent thrombosis immediately after PCI, the mechanism of this healing response during the follow-up period was uncertain because we did not perform OCT imaging at follow-up. There are several possible explanations. First, stent malapposition caused by calcium crack formation may have influenced the drug delivery of the DES. In the present study, the malapposed area was positively correlated with crack formation. Therefore, it is possible that the drug may have distributed insufficiently in the malapposed segment, causing greater neointimal proliferation and greater lumen loss at follow-up. Second, crack formation itself might be a marker of the severity of calcification. Indeed, in the present study, the calcium arc was greater in the crack group than in the no-crack group. A pathologic study has demonstrated that the prevalence of malapposed struts was significantly higher with severe calcification than with non-severe calcification. Severe calcification was reported to cause stent polymer damage (26), resulting in an unfavorable effect on drug delivery. Moreover, a recent pathologic study of a cadaver that underwent stent implantation showed that the severe medial tear was observed more frequently in the lesion with severe calcification than in those with non-severe calcification, and the amount of intimal hyperplasia was the highest in arteries with medial tears adjacent to the surface calcification (27). Another pathologic study demonstrated that severe calcification was associated with greater separation of stent struts and exaggerated intimal hyperplasia, especially in segments with medial disruption (28). Motofuji et al. reported that, among heavily calcified lesions requiring RA, nodular calcification in the culprit lesion was associated with worse long-term clinical outcomes than those without, even if favorable results after PCI were obtained (29). Our result was in accordance with the ROTAXUS trial, which evaluated the effect of RA for the calcified lesions. This randomized trial revealed that routine lesion preparation using RA did not reduce the late lumen loss of DES at 9 months, despite an initially higher acute lumen gain (30). Calcified nodules and RA theoretically increase malapposition after stent implantation, explaining poor clinical outcomes and/or greater lumen loss during follow-up. In the complex calcified coronary lesions, acute stent expansion may not always translate into the reduction of future events, even with the new generation DES. Future studies are anticipated to explore the efficacy of new modalities those are associated with favorable short- and long-term outcomes while reducing excessive healing responses after PCI injury.

## Limitation

This study has several limitations. First, we enrolled 268 patients; however, more than 30% of patients were excluded from the OCT analysis mainly due to complexity of the lesion. Second, serial change of the same cross-sectional segment at the MLA site before ballooning was evaluated as lesion-based analysis. Morphological change by ballooning was non-uniform within the lesion and the MLA site may not represent the entire lesion. Finally, OCT imaging at follow-up was not performed in this study; therefore, the precise mechanism of vessel healing remained unclear. Large-scale randomized studies need to be conducted to evaluate the impact of OCT-guided PCI on long term clinical outcomes in patients with moderate to severe lesion calcification.

## Conclusion

The OCT-guided PCI strategy demonstrated acceptable acute and 1-year clinical outcomes. The presence of calcium cracks after balloon angioplasty impact stent expansion after PCI in calcified lesions. However, its long-term efficacy could be attenuated by greater late lumen loss at 10-month follow-up.

## Data Availability

Raw data were generated at Yokohama City University Medical Center. Derived data supporting the findings of this study are available from the corresponding author on request.

## Acknowledgement

Clinical Investigators of the OCT-CALC registry Investigators were: Hiroshi Ohira (Edogawa Hospital), Tomohiko Shigemasa (International University of Health and Welfare Atami Hospital), Akiyoshi Miyazawa (Iwatsuki Minami Hospital), Ki Seok Kim (Jeju National University), Kazuki Fukui (Kanagawa Cardiovascular and Respiratory Center), Noritaka Toda (Nagatsuda Kousei General Hospital), Yukiko Morita (National Hospital Organization Sagamihara National Hospital), Jun Okuda (Omori Red Cross Hospital), Tsutomu Endo (Saiseikai Yokohamashi Nanbu Hospital), Tetsuya Tobaru and Itaru Takamizawa (Sakakibara Heart Institute), Kazuhiro Ashida (Seirei Yokohama Hospital), Kohei Wakabayashi (Showa University Koto Toyosu Hospital), Ken Kozuma (Teikyo University Hospital), Gaku Nakazawa and Yuji Ikari (Tokai University Hospital), Takahiko Kiyooka (Tokai University Oiso Hospital), Takashi Ashikaga and Taishi Yonetsu (Tokyo Medical and Dental University), Akihiro Hata (Tokyo Metropolitan Health Medical Treatment Corporation Toshima Hospital), Hiroyuki Tanaka (Tokyo Metropolitan Tama Medical Center), Teruyasu Sugano (Yokohama City University Graduate School of Medicine), Kiyoshi Hibi (Yokohama City University Medical Center), Ichiro Michishita (Yokohama Sakae Kyosai Hospital)

## Abbreviations

PCI: percutaneous coronary intervention
DES: drug eluting stent
IVUS: intravascular ultrasound
OCT: optical coherence tomography
RA: rotational atherectomy
QCA: quantitative coronary angiography
MLA: minimum lumen area
TVF: target vessel failure
ROC: receiver-operating characteristics
MSA: minimum stent area

---

**Supplement Table.**
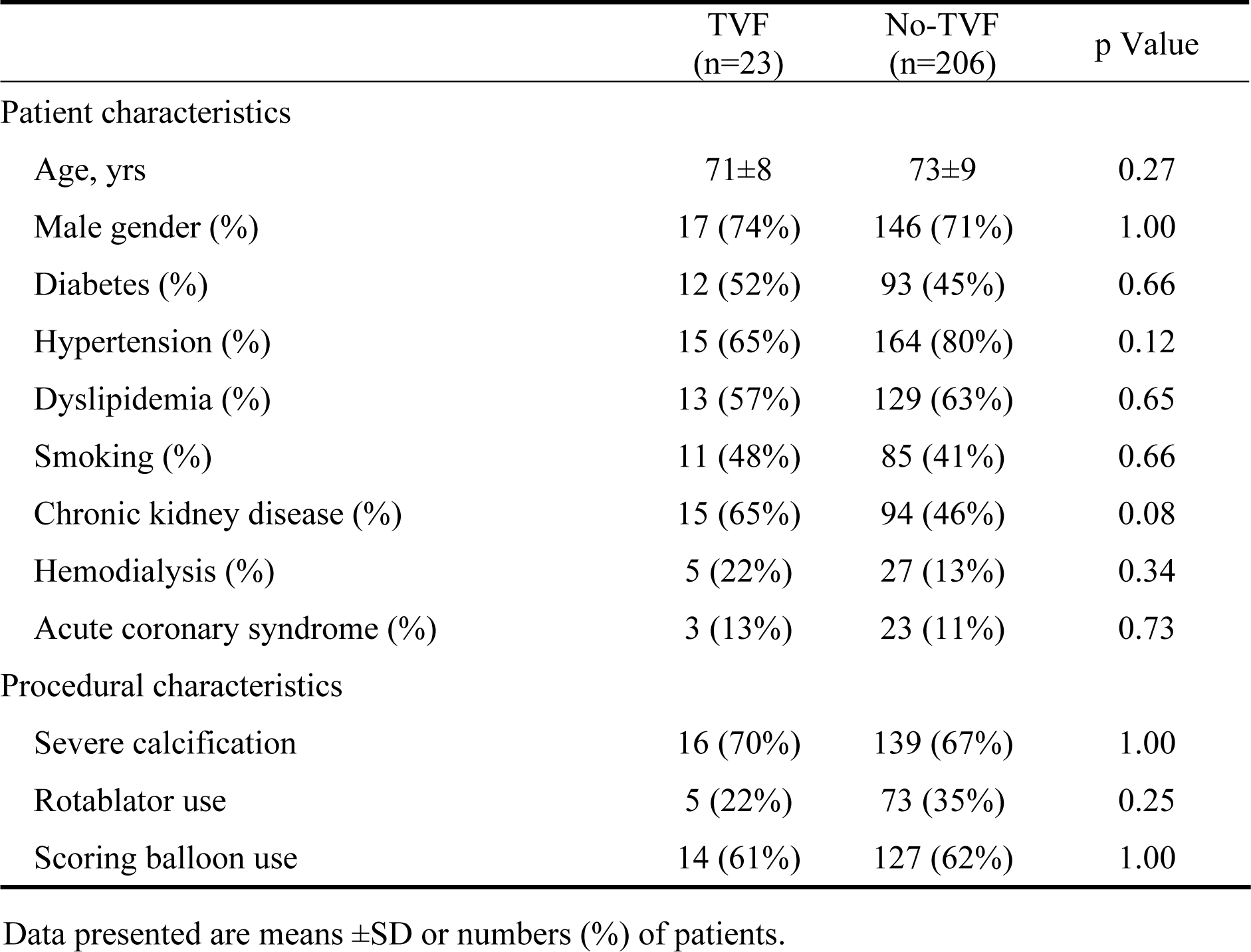
Patients and procedural characteristics between TVF and No-TVF.

Data presented are means ±SD or numbers (%) of patients. Supplement Table. Patients and procedural characteristics between TVF and No-TVF
|  | TVF<br>(n=23) | No-TVF<br>(n=206) | p Value |
| --- | --- | --- | --- |
| Patient characteristics |  |  |  |
| Age, yrs | 71±8 | 73±9 | 0.27 |
| Male gender (%) | 17 (74%) | 146 (71%) | 1.00 |
| Diabetes (%) | 12 (52%) | 93 (45%) | 0.66 |
| Hypertension (%) | 15 (65%) | 164 (80%) | 0.12 |
| Dyslipidemia (%) | 13 (57%) | 129 (63%) | 0.65 |
| Smoking (%) | 11 (48%) | 85 (41%) | 0.66 |
| Chronic kidney disease (%) | 15 (65%) | 94 (46%) | 0.08 |
| Hemodialysis (%) | 5 (22%) | 27 (13%) | 0.34 |
| Acute coronary syndrome (%) | 3 (13%) | 23 (11%) | 0.73 |
| Procedural characteristics |  |  |  |
| Severe calcification | 16 (70%) | 139 (67%) | 1.00 |
| Rotablator use | 5 (22%) | 73 (35%) | 0.25 |
| Scoring balloon use | 14 (61%) | 127 (62%) | 1.00 |

Data presented are means ±SD or numbers (%) of patients.

